# Hilbert-Envelope Features for Cardiac Disease Classification from Noisy Phonocardiograms

**DOI:** 10.1101/2020.11.17.20233064

**Authors:** Nusrat Binta Nizam, Shoyad Ibn Sabur Khan Nuhash, Taufiq Hasan

**Affiliations:** mHealth Lab, Department of Biomedical Engineering, Bangladesh University of Engineering and Technology (BUET), Dhaka 1205, Bangladesh

**Keywords:** Heart sound classification, Phonocardiogram (PCG) analysis, Hilbert envelope features

## Abstract

Cardiovascular disease (CVD) is considered a significant public health concern around the world. Automated early diagnostic tools for CVDs can provide substantial benefits, especially in low-resource countries. In this study, we propose a time-domain Hilbert envelope feature (HEF) extraction scheme that can effectively distinguish among different cardiac anomalies from heart sounds even in highly noisy recordings. The method is motivated by how a cardiologist listens to the heart murmur configuration, e.g., the intensity of the heart sound envelope over a cardiac cycle. The proposed feature is invariant to the heart rate, the position of the first and second heart sounds, and robust in extracting the murmur configuration pattern in the presence of respiratory noise. Experimental evaluations are performed compared to two different state-of-the-art methods in the presence of respiratory noise with signal-to-noise ratio (SNR) values ranging from 0-15dB. The proposed HEF, fused with standard acoustic and Resnet features, yields an average accuracy, sensitivity, specificity, and F1-score of, 94.78%(*±*2.63), 87.48%(*±*6.07), 96.87%(*±*1.51) and 87.47%(*±*5.94), respectively, while using a random forest (RF) classifier. Compared to the best-performing baseline model, this feature-fusion scheme provides a significant performance improvement (*p <* 0.05), notably achieving an absolute improvement of 6.16% in averaged sensitivity. In the case of 0dB SNR, the proposed feature alone provides a 9.2% absolute improvement in sensitivity compared to the top baseline system demonstrating the robustness of the HEF. The developed methods can significantly impact computer-aided auscultation (CAA) systems when deployed in noisy conditions, especially in low-resource settings.

## I. Introduction

Cardiovascular diseases (CVDs), the leading cause of morbidity in the world, claims an estimated 17.9 million lives every year worldwide while it contributes to the highest percentage of deaths, 30%, among non-communicable diseases (NCDs) [1], [2]. The prevalence of CVDs is also highest in low and middle-income countries where the weighted pooled prevalence of CVDs is 5% [3], [4]. Although coronary heart diseases are the most frequently occurring CVD, valvular heart diseases (VHDs) contribute significantly to the overall morbidity and mortality. VHDs are present in 17% of the community with heart failure in the US [5]. The US has a 2.5% prevalence of significant VHDs [6], and about 10% to 20% of cardiac surgical procedures contribute to VHDs [7]. In many developing countries, VHDs have an additional burden due to rheumatic fever, which can cause progressive valve inflammation and fibrosis, leading to rheumatic heart diseases (RHDs) [8]. Overall, VHDs are of major health concern for physicians and general people across the globe.

Early detection can drastically reduce the complications of VHDs. Although echocardiography is the gold standard diagnostic tool for VHDs, cardiac auscultation is perhaps the most widely used and cost-effective procedure for screening VHDs. Procedures such as cardiovascular magnetic resonance imaging and exercise testing are also used for diagnosing VHDs [9]. However, particularly in low-resource settings, the effective use of cardiac auscultation is vital for early diagnosis of VHDs. The shortage of cardiologists and trained physicians represents another major challenge in the underserved communities [10]. In these circumstances, machine learning-based automated VHD detection systems can have a significant impact, particularly systems implemented using smartphone applications or using embedded systems within smart digital stethoscopes [11]. However, noise robustness is an important factor for automated PCG analysis in low-resource settings.

Various methods have been proposed in the past for automated detection and classification of PCGs acquired via a digital stethoscope. PCG signal analysis generally involves segmentation, feature extraction and finally classification. Important characteristics of heart sounds are interpretable from representing them in different domains [12]. Time-domain features contain temporal and statistical information about the various cardiac events [13]. Frequency domain features use band-pass filter banks, zero-crossing analysis [13]–[15]. However, a majority of previous studies have considered features extracted from time-frequency domain representations, such as the S-transform [16], short-time Fourier transform (STFT) [17], Wigner-Ville distribution (WVD) [18], [19], Choi–Williams distribution (CWD) [20], [21], wavelet transform [22]–[24], short-time modified Hilbert transform (STMHT) [25], and mel-scaled wavelet transform [13], [26], [27]. Envelope based features [28], mel-frequency cepstral coefficients (MFCCs) [29]–[32] and fractal features [33], [34] have also been used in analyzing PCGs [13]. Support vector machine (SVM) [35], random forest (RF) [36], k-Nearest Neighbor (k-NN) [37], multilayer perceptron (MLP) [30], [38] are some of the commonly used classification algorithms used in PCG analysis. In recent studies, deep learning algorithms such as 1D and 2D convolutional neural networks (CNN) are being used for this task [39]–[41]. However, only a few of the previous studies have considered the analysis of the murmur configuration patterns frequently used by cardiologists during auscultation.

In this work, we have developed a Hilbert envelope based feature set that delineates the intensity variation of the heart sounds, known as murmur configuration, effectively used by cardiologists during auscultation. These configurations define the increasing (crescendo) or decreasing (decrescendo) nature of the murmur while also indicate their temporal relationship with the first and second heart sounds. To evaluate the robustness of the proposed method, we contaminate the PCG signals by mixing respiratory sounds [42] with the recordings to simulate real-life clinical conditions at different noise levels.

This paper is organized as follows. In Sec. II, we discuss the motivations behind our proposed method. In Sec. III, we describe our proposed feature extraction scheme, followed by Sec. IV containing descriptions of the baseline systems. In Sec. V, the datasets used are described. Section VI details the evaluation of the proposed features in comparison with the baseline systems. In Sec. VII, we discuss the limitations and future directions of our study, and finally, summarize our findings in Sec. VIII.

## II Background

In this section, we review the origin of heart sounds and their different physiological and pathological characteristics. We highlight some major valvular heart diseases with a particular focus on how a physician interprets these sounds.

### A. Heart sound fundamentals

Auscultation of the heart sound (i.e., PCG) is an essential procedure for the initial screening and diagnosis of different CVDs. PCG signal is produced by the contraction of the heart, valves, and vessels, which is captured using a stethoscope. The functional heart mainly produces two heart sounds: the first heart sound (*S*_1_) and second heart sound (*S*_2_). *S*_1_ is heard at the beginning of systole, which is associated with the closure of mitral and tricuspid heart valves. At this time, increasing pressures within the ventricles decelerate the moving valve leaflets and cord structures. Ventricular pressure rapidly drops at the end of ventricular systole, which causes the backflow of blood from the aorta and pulmonary artery. These events cause the closure of the aortic and pulmonic valves, producing the *S*_2_ sound. Heart sounds contain six different characteristics analyzed by the physician for a complete assessment. These are intensity, duration, pitch, quality, and timing [43], [44].

### B. Overview of heart murmurs

Heart murmur sounds are mainly generated because of turbulent blood flow. In normal conditions, the blood flow through the vascular bed is laminar, smooth, and silent. As a result of hemodynamic or structural changes, laminar flow can become disturbed, which in turn produces audible noise. In general, murmur sounds can be generated because of the following reasons:

- Blood flow across a partial obstruction
- Increased blood flow across normal structures
- Ejection of blood into a dilated chamber
- Regurgitant flow across an inadequate valve
- Abnormal shunting of blood from one vascular chamber to another because of a lower pressure gradient

Physicians generally quantify the intensity of the murmurs by using a predefined grading scheme. Grading schemes for the systolic and diastolic murmurs are shown in Table I.

**TABLE I:**
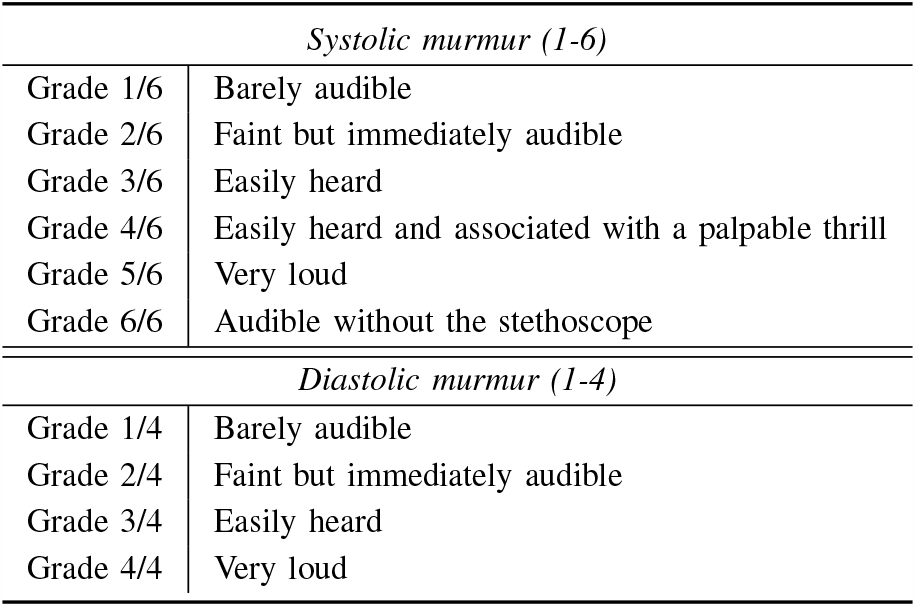
Intensity grading scheme used by physicians for systolic and diastolic murmurs

The configuration of a murmur refers to the shape of its waveform as observed on a PCG signal. This configuration is determined by the intensity and pressure gradients of vascular flow during systole or diastole. The shape of a murmur mainly describes the intensity change from the beginning until the end within a heart cycle. A crescendo type murmur gradually increases intensity with the increase of pressure gradient. On the other hand, a decrescendo murmur decreases its intensity with the pressure gradient decline. In a crescendo-decrescendo murmur, also referred to as the diamond-shaped murmur, the intensity, and pressure gradient first increase then decrease. The plateau-shaped murmur is of the same intensity throughout its duration. Different types of murmur patterns for various valvular heart diseases are illustrated in Table II. Murmurs are heard the loudest in the region where a particular valve is affected. Mainly there are four anatomical sites where auscultation is performed for listening to murmurs. These are the Aortic area, Pulmonary area, Tricuspid area, and Mitral area.

**TABLE II:**
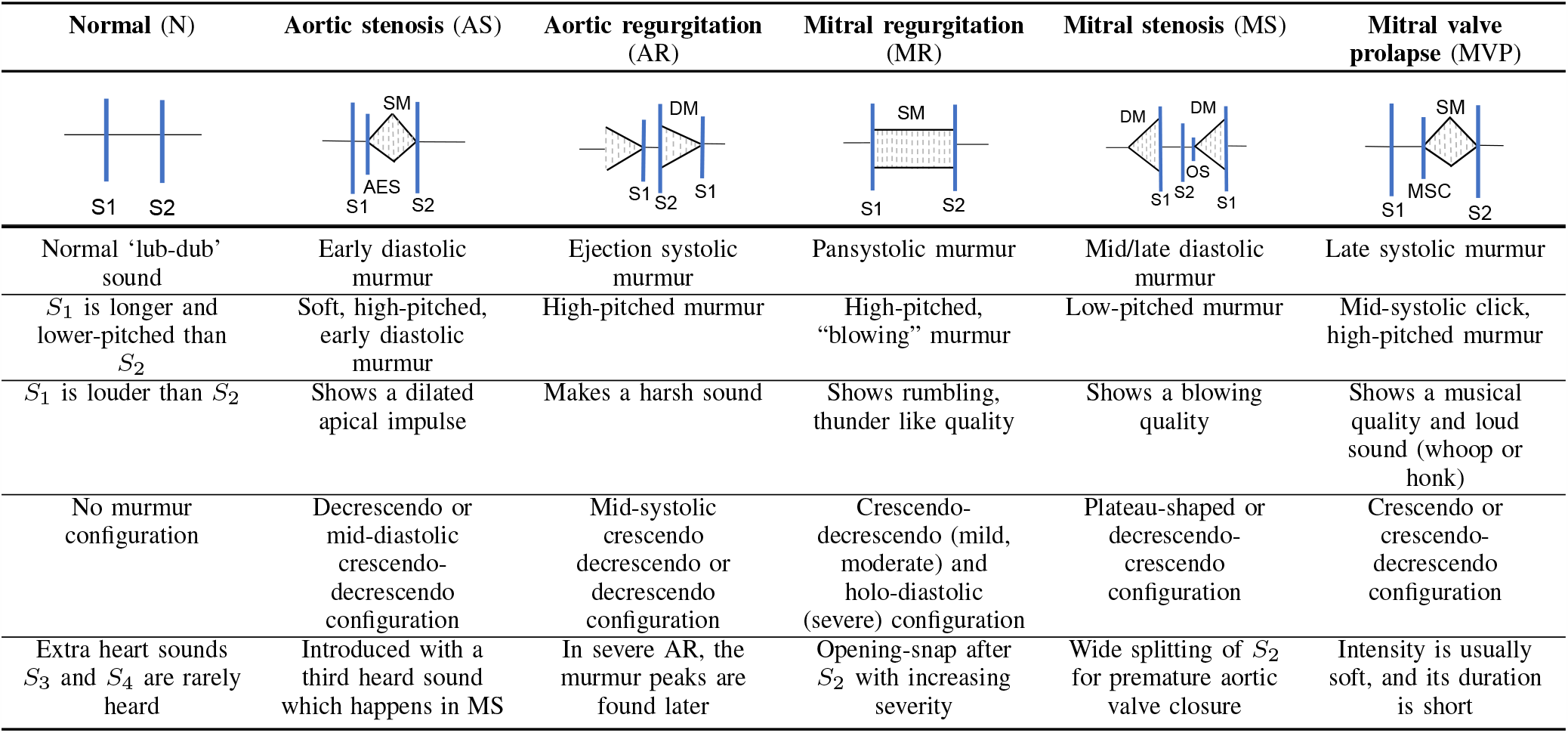
Heart sound characteristics for different VHDs illustrating different murmur patterns

| Normal (N) | Aortic stenosis (AS) | Aortic regurgitation (AR) | Mitral regurgitation (MR) | Mitral stenosis (MS) | Mitral valve prolapse (MVP) |
| --- | --- | --- | --- | --- | --- |
| Normal ‘lub-dub’ sound | Early diastolic murmur | Ejection systolic murmur | Pansystolic murmur | Mid/late diastolic murmur | Late systolic murmur |
| $S_1$ is longer and lower-pitched than $S_2$ | Soft, high-pitched, early diastolic murmur | High-pitched murmur | High-pitched, “blowing” murmur | Low-pitched murmur | Mid-systolic click, high-pitched murmur |
| $S_1$ is louder than $S_2$ | Shows a dilated apical impulse | Makes a harsh sound | Shows rumbling, thunder like quality | Shows a blowing quality | Shows a musical quality and loud sound (whoop or honk) |
| No murmur configuration | Decrescendo or mid-diastolic crescendo-decrescendo configuration | Mid-systolic crescendo decrescendo or decrescendo configuration | Crescendo-decrescendo (mild, moderate) and holo-diastolic (severe) configuration | Plateau-shaped or decrescendo-crescendo configuration | Crescendo or crescendo-decrescendo configuration |
| Extra heart sounds $S_3$ and $S_4$ are rarely heard | Introduced with a third heard sound which happens in MS | In severe AR, the murmur peaks are found later | Opening-snap after $S_2$ with increasing severity | Wide splitting of $S_2$ for premature aortic valve closure | Intensity is usually soft, and its duration is short |

Murmurs can be classified broadly as systolic or diastolic. All systolic murmurs occur during ventricular systole between the first and second heart sounds. Diastolic murmurs can be heard between the second and first heart sounds. Murmur patterns are different for different valvular heart diseases. Valvular heart diseases can be identified from the PCG and murmur patterns [44], [45]. A comparative analysis of murmurs observed for different VHDs are provided in Table II [43], [44]. These diseases are described in the following sub-sections.

1. *Aortic stenosis (AS):* The most likely cause of this systolic murmur is age-related calcification of the aortic valve. These calcified changes in the heart valve can also lead to valve deformation. About 1%-2% of the world population are born with an abnormal bicuspid aortic valve [45].
2. *Aortic regurgitation (AR):* Aortic regurgitation is an early diastolic murmur, which is also known as aortic insufficiency. It can be caused by rheumatic heart disease, Marfan syndrome, osteogenesis imperfecta, congenital bicuspid valve, or dissecting aortic aneurysm. The leakage of the prosthetic aortic valve can also cause this abnormality [43]. This murmur is accompanied by a systolic ejection murmur (SEM) produced by the increase of the left ventricular stroke volume.
3. *Mitral regurgitation (MR):* Mitral regurgitation murmur, also known as mitral insufficiency, is caused by abnormalities in the mitral valve leaflet. Increased pressure at the aortic valve creates regurgitation of blood through the mitral valve producing this murmur. This murmur can be heard at early systole, mid-systole, or late systole and may be holosystolic (beginning at *S*_1_ and continuing to *S*_2_).
4. *Mitral stenosis (MS):* Mitral stenosis is caused due to congenital defects or chronic rheumatic valvulitis. Valvulitis leads to scarring and calcification of the valve, adversely affecting its normal functionalities. At the beginning of this murmur, an opening snap is heard after the closure of the aortic valve and iso-volumetric relaxation. This murmur is produced as the turbulent blood rapidly flows through a rigid, narrowed mitral valve opening. The increase of turbulence results in its characteristic pre-systolic crescendo. The duration and other murmur characteristics may vary with the degree with disease severity (mild, moderate, or severe).
5. *Mitral valve prolapse (MVP):* MVP murmur appearing mainly in late systole is caused by the prolapsed mitral valve. In some cases, it is isolated or accompanied by a non-ejection mid-systolic click (MSC) caused by the ballooning up into the left atrium. MVP is also known as the click murmur syndrome.

### C. Motivation for envelope-based features

In real-world scenarios, heart sounds recorded using a digital stethoscope can be corrupted by different noises, most commonly the patient’s respiration sounds. The PCG signal by itself is non-stationary with various nuances, as discussed. However, the PCG signal envelope incorporates the simple amplitude patterns that carry relevant information regarding the underlying abnormalities. The physicians are also trained to identify these murmur sounds based on their configuration (e.g., crescendo, decrescendo, crescendo-decrescendo), as described in the previous sub-sections. We hypothesize that the envelope of a PCG signal can effectively represent the murmur configuration and can thus be used to design effective features to distinguish the nuances of the PCG signal observed in different VHDs. Since the respiratory sound is relatively stationary compared to the heart sound, the murmur patterns can be more effectively extracted from the signal envelopes for this particular type of noise.

## III. Hilbert-Envelope Feature (HEF) Set

This section describes the steps performed to extract the proposed Hilbert-envelope feature (HEF) set to be used for VHD classification. The process is represented in the block diagram of Fig. 1.

**Fig 1:**
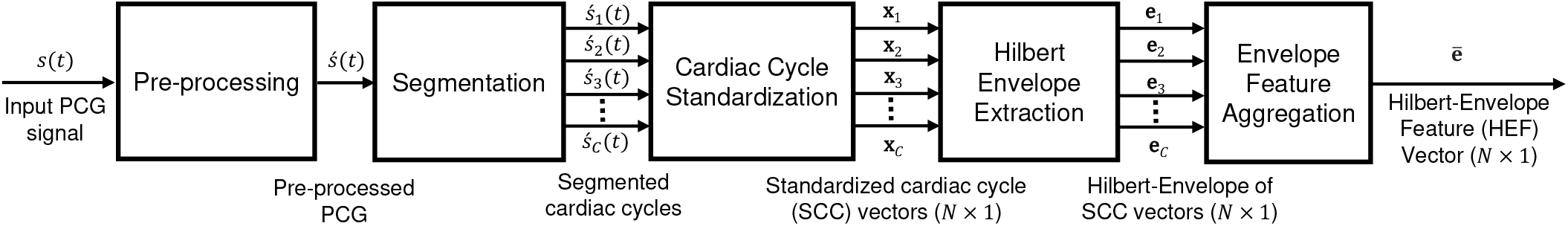
A block-diagram showing the various steps in extracting the proposed Hilbert-envelope feature (HEF) set.

### A. Pre-processing

Each PCG signal is first down-sampled to 1000 Hz. Next, it is band-pass filtered in the frequency range 25–400 Hz using two cascaded second-order Butterworth filters (low-pass and high-pass). This is done since the fundamental heart sounds, *S*_1_ and *S*_2_, generally lie in the frequency range of 10 − 250 Hz while pathological murmurs typically have frequency components below 600 Hz [12]. The filtered signal is then processed using the spike removal algorithm in [46]. In the block diagram of Fig. 1, the original PCG signal is denoted by *s*(*t*) and the pre-processed signal by *s ′* (*t*).

### B. Segmentation

In this work, we use a logistic regression-hidden semi-Markov model (HSMM) based segmentation method presented in [47] to identify the *S*_1_ and *S*_2_ locations. This segmentation method incorporates logistic regression (LR) into a duration dependant HSMM for emission probability estimation and an extended Viterbi algorithm for decoding the most likely sequence of heart sound states [46]. Assuming there are *C* cardiac cycles, *s′* (*t*) is divided into cardiac cycle signals 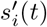, where *i* = 1 *…C*. All partial cardiac cycles are discarded.

### C. Extraction of standard cardiac cycles (SCC)

As discussed in Sec. II and summarized in Table II, the pathological murmur sounds show characteristic patterns in the signal envelope (e.g., crescendo, decrescendo). However, these patterns are only meaningful when observed within a full cardiac cycle. Accordingly, after performing segmentation to extract the cardiac cycles from each PCG recording, each cardiac cycle is down-sampled to a fixed *N* dimensional vector, such that the heart sound states *S*_1_, systole, *S*_2_ and diastole, consist of 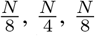, and 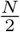 samples respectively. This *N ×* 1 down-sampled vector is referred to as the *standardized cardiac cycle* (SCC) vector and we denote it by **x**_*i*_ extracted from the *i*-th cardiac cycle of *s*(*t*). This standardization process removes the variability within different cardiac cycles due to differences in heart-rate and allows for a systematic comparison between cardiac cycles obtained from different recordings.

### D. Hilbert-envelope extraction

In this step, the discrete Hilbert transform is applied to the sequence of SCC vectors **x**_*i*_ where *i* = 1 *…C* is the cardiac cycle index. Denoting the *n*-th component of the SCC vector **x**_*i*_ as *x*_*i*_(*n*), its Hilbert transform is given by

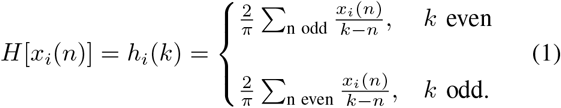

The complex analytic signal *A*[*x*_*i*_(*n*)] is then computed as

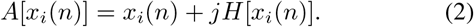

The absolute value of *A*[*x*_*i*_(*n*)] represents the envelope signal, *e*_*i*_(*n*) given by

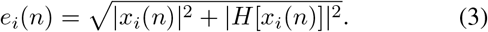

Fig. 2 shows the PCG signal in various stages of the envelope extraction process for a MVP recording at 15db SNR.

**Fig 2:**
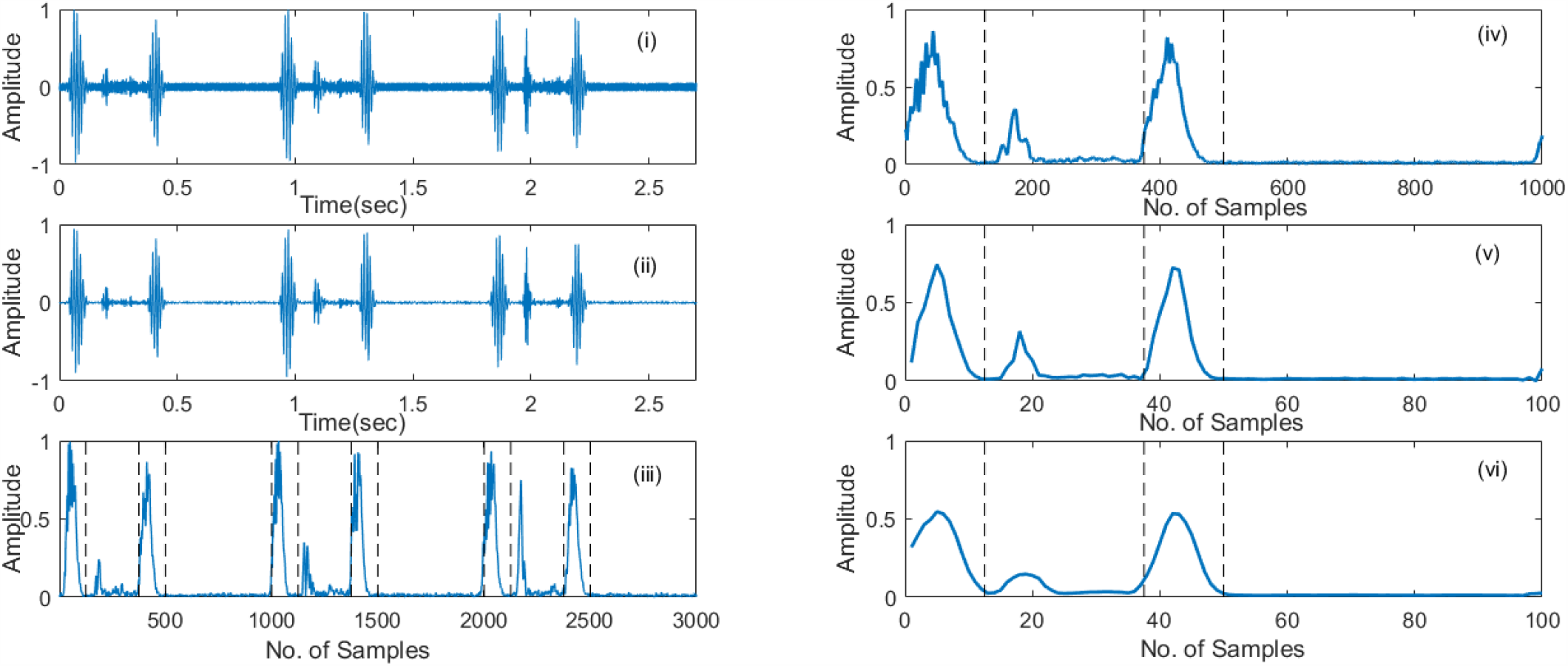
A PCG signal in various stages of the proposed envelope feature extraction procedure. (i) 15db noisy PCG signal, (ii) pre-processed PCG signal, (iii) standardized Hilbert envelope, (iv) after first part of feature aggregation process, averaged envelope, (v) and (vi) after later parts of feature aggregation process, downsampled and smoothed envelope respectively.

### E. Envelope feature aggregation

The envelope signal extracted from the *i*-th cardiac cycle can be represented as a vector **e**_*i*_ where *i* = 1 … *C*. Each envelope signal is then amplitude normalized by its maximum value. The number of cardiac cycles may vary from subject to subject even for a fixed length of PCG signal. To account for this variability, the envelopes extracted from *C* cardiac cycles are averaged to extract the Hilbert Envelope feature (HEF) vector as

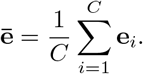

The HEF vctor ē is of dimension *N* as defined by the SCC extraction step. In our work, we use the value of *N* = 1000. As the final steps, the HEF vectors ē are further downsampled for dimensionality reduction and smoothed using a moving average (MA) filter.

## IV. System description

This section describes the two baseline systems used for a comparative study and discusses the implementation details of the proposed heart sound classification system.

### A. Baseline-1: MFCC-DWT-SVM system

This baseline architecture follows the top-scoring algorithm presented in [31] where the dataset [32] was introduced. This system uses fused MFCC and DWT features as input to an SVM classifier. 19 dimensional MFCC features are extracted using 240 sample frames with a step-size of 80, while 24 dimensional DWT features are extracted using 4000 sample frames with a step-size of 1000. Sampling frequency were 8000 Hz for both MFCC and DWT. Although the authors reported 97.9% accuracy, with the parameters set as described in [31] and using the provided code by the authors [32], we obtained an average accuracy of 94.84%. This method will be referred to as Baseline-1 in the remainder of this paper.

### B. Baseline-2: ResNet-IS09-RF system

This system follows [48] where the ResNet architecture and the openSMILE toolkit are used for audio/acoustic feature extraction while a random forest (RF) is used for classification. The feature sets are described below.

1. *Spectro-temporal ResNet feature set:* Short-term Fourier power spectrum is used to extract spectrogram images from the input PCG signal. The spectrogram images are then passed into a ResNet-50 model [49] pre-trained on the ImageNet dataset to extract the high-dimensional features. This results in a feature vector of dimension 100352.
2. *IS09 acoustic feature set:* The acoustic feature set used is described in [50], which is commonly known as the INTERSPEECH 2009 Emotion Challenge (IS09) feature set. This feature set includes 16 low-level descriptors (LLD) and 12 of their functionals that result in a total of 384 dimensional feature vector. Table III summarizes the LLD and the functionals used [50].

**TABLE III:** Low-level descriptors (LLD) and functionals used to extract the IS09 acoustic feature set.

| LLD (16 × 2) | Functionals (12) |
| --- | --- |
| Zero-crossing-rate | Mean |
| RMS Energy | Standard Deviation (std) |
| Pitch energy (F0) | Kurtosis, Skewness |
| Harmonics-to-noise ratio | Extremes: value, rel. position, range |
| MFCC 1-12 | Linear Regression: offset, slope, MSE |

### C. Proposed HEF-based systems

The proposed HEF feature set is utilized to develop three different system configurations for the VHD classification task. These configurations are (i) HEF-only system set with random forest classifier (HEF-RF), (ii) a combination of HEF, ResNet and IS09 features with the RF classifier (HEF-ResNet-IS09-RF), and (iii) a combination of HEF, ResNet and IS09 features with the support vector classifier (HEF-ResNet-IS09-SVC). The overall flow diagram of the proposed configuration (ii) and (iii) are shown in Fig. 3.

**Fig 3:**
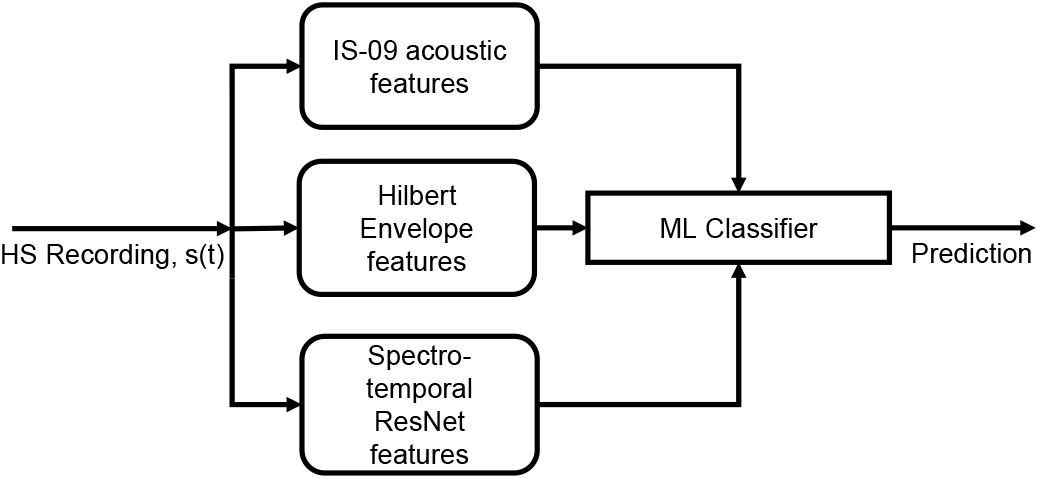
Block diagram of the proposed system configurations.

## V. Data sources

### A. Open-access PCG dataset

In this work, we utilize the PCG recordings made available in [32]. This dataset contains 1000 recordings from five categories, namely, AS, MR, MS, MVP and normal, each containing 200 recordings. Each PCG recording in the dataset contains 3 complete cardiac cycles of noise free heart sound. The PCG signals in this dataset are sampled at 8kHz.

### B. Preparation of a noisy dataset

Evaluating the noise-robustness of our algorithms requires access to noisy multi-class PCG datasets. Since such data is not available, we degraded the heart sounds with lung sounds as they are the most frequently encountered noise in cardiac auscultation. We use the respiratory sound dataset reported in [42] as noise. This dataset consists of a total of 920 lung sound recordings collected from 126 patients.

For a clean PCG signal, the following steps are performed for generating its noisy version. A respiratory sound signal is selected randomly. Since the length of the PCGs are much shorter (2 − 3 seconds) compared to the respiratory sounds (20 seconds), these segments (noise) are clipped at random positions to match the length of the PCG (clean signal). Amplitude of noisy signals is adjusted according to the desired SNR defined by

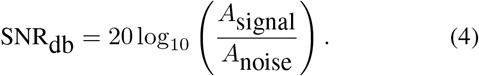

The resulting noisy signal is amplitude normalized.

### C. Dataset splitting and cross-validation

We organize the open-access dataset [32] to perform five-fold cross-validation experiments for clean and noisy conditions. The full clean dataset is first divided into five equal folds. One set is used for each fold for testing, while the remaining sets are used for training. For the noisy experiments, the same PCG data folds are used for the cross-validation with the addition of the corresponding noisy PCG recordings. We ensure that the same noise segment (respiratory sound) is never used for both training and testing for any fold.

## VI. Experimental evaluation

The performance of the proposed systems are evaluated and compared with the two baseline systems using the datasets described in Sec. V. We systematically perform experiments first to analyze and select the HEF parameters. Afterward, we perform the evaluations in clean and noisy conditions.

### A. Parameter selection for HEF extraction

In this experiment, optimal parameters for extracting the HEF set are determined. The HEF extraction includes two parameters (i) feature dimension and (ii) length of the MA smoothing filter. While discerning the optimal value of one parameter, the value of all other parameters is kept unchanged.

In the first experiment, we observe the system performance on the 5-fold cross-validation task described in Sec. V-C by varying the HEF size from 100 to 1000 with an increment of 100 without smoothing. Both SVC and RF classifiers are used. We use sensitivity as the discerning performance metric. The results are summarized in Fig. 4, where we observe that the optimal HEF size can be found at 100 and 500 for the RF and SVM classifiers, respectively.

**Fig 4:**
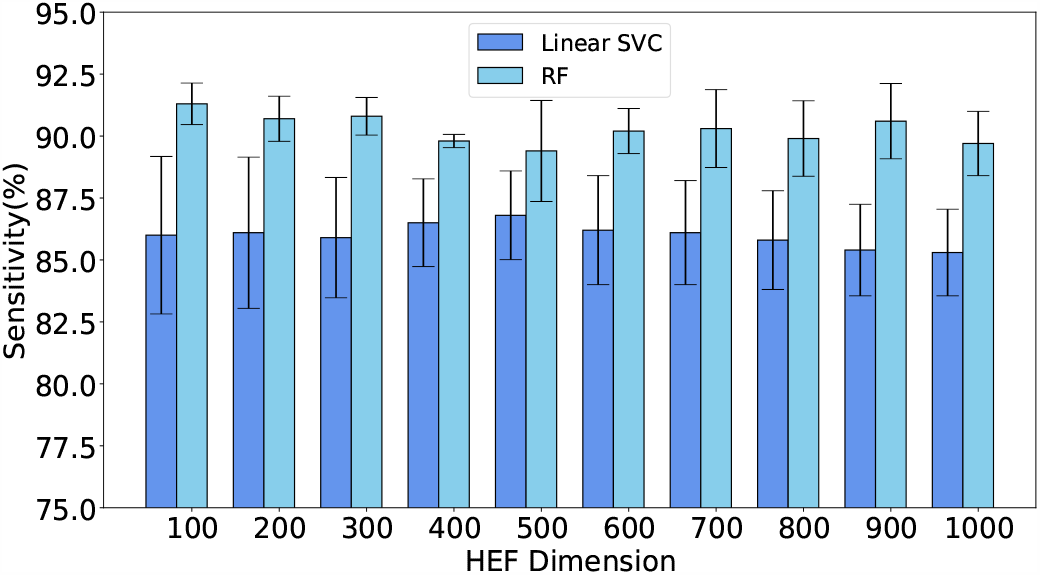
Effect of HEF dimension on sensitivity. Error bars represent one standard deviation of uncertainty

In the next experiment, the HEF sizes of 100 and 500 are smoothed by applying MA filters of length 2 to 10 with an increment of 1. The results depicted in Fig. 5 show that the optimal sensitivity of 92.7% is obtained for HEF size of 100 with a MA filter of length 6 while using the RF classifier. Thus, for the remaining experiments we set the HEF size to 100 and MA filter length to 6.

**Fig 5:**
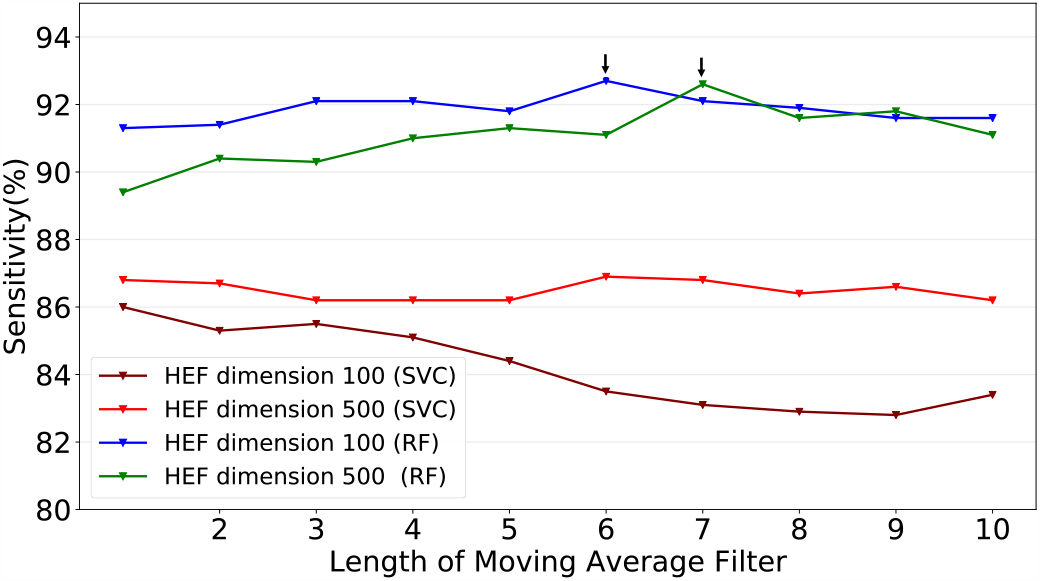
Effect of envelope smoothing on sensitivity. Arrows mark the best performing points of the plot.

### B. Results on the clean condition

In this experiment, five-fold cross-validation is performed on the original clean recordings of the open-access dataset [32]. In Table IV, performance metrics of the proposed systems are compared with the baseline systems. The results show that the proposed HEF-ResNet-IS09 systems show the best overall performance compared to the baseline systems with respect to the accuracy, sensitivity, specificity, and F-1 score. Since all systems perform near the 90% accuracy mark in the clean condition, we focus our attention on the noisy conditions discussed in the next section.

**TABLE IV:** Experimental results on clean PCG dataset

| System | Accuracy (%) | Sensitivity (%) | Specificity (%) | F1-Score (%) |
| --- | --- | --- | --- | --- |
| Baseline-1:<br>MFCC-DWT-SVM | 94.84<br>($\pm 4.68$) | 87.1<br>($\pm 11.7$) | 96.78<br>($\pm 2.93$) | 87.08<br>($\pm 11.78$) |
| Baseline-2:<br>ResNet-IS09-RF | 97.43<br>($\pm 1.74$) | 93.2<br>($\pm 4.17$) | 97.99<br>($\pm 0.93$) | 93.4<br>($\pm 4.22$) |
| HEF-RF | 93.48<br>($\pm 4.07$) | 83.70<br>($\pm 10.17$) | 95.92<br>($\pm 2.54$) | 82.99<br>($\pm 11.15$) |
| HEF-ResNet-IS09-SVC | **98.84**<br>($\pm 0.45$) | **97.00**<br>($\pm 1.26$) | **99.27**<br>($\pm 0.28$) | **97.11**<br>($\pm 1.11$) |
| HEF-ResNet-IS09-RF | 98.58<br>($\pm 0.34$) | 96.50<br>($\pm 0.89$) | 99.10<br>($\pm 0.22$) | 96.39<br>($\pm 0.87$) |

### C. Results on noisy conditions

In this experiment, the performance of the baseline and proposed systems for PCG signal classification are evaluated in presence of respiratory noise. Again, a five-fold cross-validation scheme is used on recordings from datasets described in Sec. V where train and test sets contain both clean and noisy recordings. The experimental results comparing the baseline, and proposed systems in the noisy conditions are summarized in Fig. 6 and 7. In Fig. 6, the performance metrics are obtained for a mixed test condition that includes clean and noisy PCG (including 0 − 15dB SNR). From the results, we observe that in this mixed noisy condition, the proposed HEF-ResNet-IS09-RF systems significantly outperform both the baseline systems. The system yields an average accuracy, sensitivity, specificity, and F1-score of, 94.78% (*±* 2.63), 87.48%(*±* 6.07), 96.87 % (*±* 1.51) and 87.47% (*±*5.94), respectively. The p-values obtained from t-tests comparing the accuracy of HEF-ResNet-IS09-RF with baseline-1 & 2 were found be significant at the 0.05 level (*p <* 0.05).

**Fig 6:**
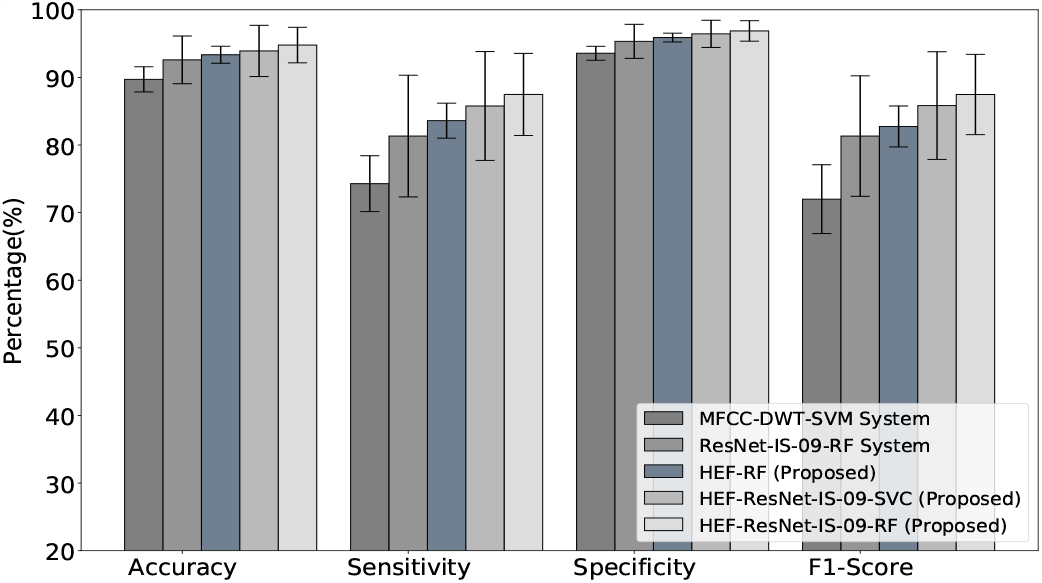
Performance evaluation of different methods on the mixed test-set including clean and noisy PCG (0 −15dB SNR).

**Fig 7:**
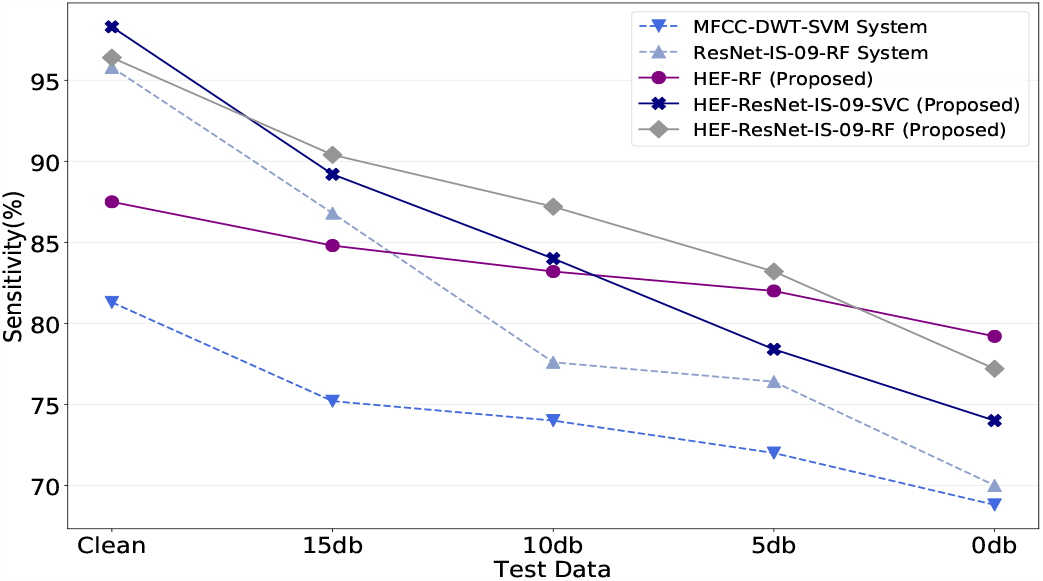
Performance evaluation of different methods with respect to sensitivity in different noise levels.

In order to observe the performance of the methods separately in different noise levels, we turn our attention to Fig. 7, where sensitivity values across different methods for each condition (Clean, 15dB, 10dB, 5dB, and 0dB SNR) are compared. In this figure, we observe that sensitivity drops as the SNR decreases for all methods. For baseline 1 and 2, although the performances in the clean condition were reasonable, sensitivities of both systems sharply decline as the noise level is increased. However, the HEF-RF system, that uses the proposed features alone, shows superior performance compared to baseline 1 for all noise levels. Notably, in the highest level of noise at 0dB SNR, the HEF-RF system provides the best sensitivity of 79.2%(*±* 6.26) showing an absolute improvement of 9.2% compared to baseline-2 system. This demonstrates the ability of the proposed HEF to encode the unique heart sound murmur characteristics embedded in the signal envelopes, even in high levels of respiratory noise. Note that all methods compared in Fig. 7 include noisy data in their training set. Thus, we conclude that the improvement observed in these results is due to the effectiveness of the proposed feature-set.

## VII. Discussions

One limitation of the proposed feature extraction method is its dependency on an accurate heart sound segmentation algorithm. If the first and second heart sounds are not correctly identified, the subsequent PCG standardization and envelope extraction process may fail. This could mean that the presence of impulsive noise due to sensor friction or environmental sounds may interfere with the method as these sounds can be misidentified. Nevertheless, we have experimentally demonstrated the effectiveness of the proposed features in the presence of respiratory noise at 0dB SNR. We will consider a further evaluation of the proposed method in real-world noisy PCG recordings.

## VIII. Conclusion

In this study, we present a novel time-domain feature set for VHD classification based on the Hilbert envelope of the heart sound. The design of the feature was motivated by methods followed by physicians during cardiac auscultation that involves analyzing the murmur configuration, e.g., intensity variation of the PCG over time. The proposed feature set, along with acoustic and ResNet features, has been shown to provide significantly improved performance compared to existing VHD classification methods in the presence of respiratory sound noise. The best performing system has provided an average accuracy of 94.78% and sensitivity of 87.48% showing an absolute improvement of 6.16% (*p <* 0.05) compared to a state-of-the-art method. The proposed feature has been found to be robust even in a 0dB SNR condition providing the highest sensitivity at 79.2%, demonstrating the effectiveness of the envelope-based approach.

## Data Availability

Datasets used in the study are available online at:
Y. Khan, Classification of heart sound signal using multiple features. [Online]. Available: https://github.com/yaseen21khan/Classification-of-Heart-Sound-Signal-Using-Multiple-Features-

